# Comparison of characteristics of bimanual coordinated movements in older adults with frailty, pre-frailty, and robust health

**DOI:** 10.1101/2024.12.02.24318299

**Authors:** Shoya Fujikawa, Shin Murata, Akio Goda, Shun Sawai, Ryosuke Yamamoto, Yusuke Shizuka, Takayuki Maru, Kotaro Nakagawa, Hideki Nakano

## Abstract

**Introduction:** Despite the growing concern regarding a potential increase in the number of older adults with frailty owing to an aging global population, the characteristics of bimanual coordination in such older adults remain unclear. This study aimed to compare bimanual coordinated movements among community-dwelling older adults with frailty, pre-frailty, and robust health and identify the specific characteristics of these movements in older adults with frailty.

**Methods:** Participants were divided into frail, pre-frail, and robust groups on the basis of Kihon Checklist scores; they performed in-phase (tapping the thumb and index finger together as fast as possible) and anti-phase (alternating the movement between the left and right fingers) bimanual coordination tasks, and the task parameters were then compared among the groups.

**Results:** The total travel distance during the anti-phase task in the frail group was significantly shorter than that in the robust group. However, all three groups showed lower finger dexterity during the anti-phase task than the in-phase task and in the left hand than in the right hand.

**Conclusions:** Older adults with frailty show less movement in bimanual coordination tasks than robust older adults, suggesting that bimanual coordination tasks may be useful tools for assessing frailty.

## 1 Introduction

The percentage of older adults in the population is increasing annually worldwide. According to the World Health Organization, between 2020 and 2050, the population of individuals aged ≥60 years is estimated to double to 2.1 billion, and the population of those aged ≥80 years is projected to triple to 426 million (1). The rapid aging of the global population has driven interest in improving the understanding of healthy aging and identifying assessment methods for it (2,3). Healthy aging is a complex multidimensional concept that encompasses biological, functional, lifestyle, and psychosocial factors (3). Additionally, achievement of healthy aging requires early interventions to prevent significant declines in physical and cognitive functions (4). The amount of older adults with frailty is also expected to increase as the older adult population grows. Frailty is characterized by a heightened risk of comorbidities and mortality (5). However, physical function in individuals with frailty has been reported to improve with appropriate interventions (6). Furthermore, prevention of frailty has been identified as a key future project in public health (7) and holds significant social importance. Therefore, establishing a method for assessing frailty is crucial for maintaining the health of older adults.

In daily life, hands are the most frequently used body part (8), and healthy older adults have been found to engage in activities involving both hands more frequently than those involving only one hand (9). Upper limb function in humans has been shown to change with age. Ingram et al. compared upper limb muscle strength, positional and superficial sensations, one-handed dexterity, bimanual coordination, muscle power stability, and functional performance in healthy participants aged 20–95 years (10). Their results showed that the participants’ performance on all the parameters decreased with age, and the decline in bimanual coordination was particularly significant. Additionally, studies have reported that bimanual movements exhibit decreased accuracy, increased variability, and prolonged motor execution times with age (11). These results indicate that bimanual coordinated movements play an important role in the daily lives of older adults, and that the coordination underlying these movements declines with age.

Although older adults with frailty have been reported to exhibit lower dexterity in one-handed movements than healthy older adults (12,13), the characteristics of bimanual coordinated movements in older adults with frailty have not been clarified. Frailty in older adults has also been reported to result in less independence in activities of daily living than healthy older adults (14), a higher risk of falling (15), and sarcopenia (16). In contrast, higher finger dexterity has been reported to be associated with better predictive postural control ability in stepping movements (17), and improved upper limb function has been reported to enhance gait ability and overall quality of life (18). Furthermore, sensory stimulation from the fingertips resulting from light contact has been shown to reduce ankle joint and body sway in the standing posture of older adults (19), suggesting that upper limb function, including finger function, can compensate for the decline in gait ability and standing balance. Therefore, if the characteristics of bimanual coordinated movements in frail older adults are clarified, they can be used as an early intervention tool to detect frailty. Thus, this study aimed to compare the bimanual coordinated movements of community-dwelling older adults with frailty, pre-frailty, and robust health and determine the characteristics of bimanual coordinated movements in older adults with frailty. We hypothesized that older adults with frailty would exhibit lower bimanual coordination than robust older adults.

## 2 Methods

### 2.1 Participants

This cross-sectional study was conducted with 358 community-dwelling older adults who participated in physical fitness assessment sessions held in two cities in September 2023. The exclusion criteria for the participants were: (i) age < 65 years; (ii) Mini-Mental State Examination (MMSE) scores < 24; (iii) presence of hand dexterity impairments due to musculoskeletal or central nervous disease; (iv) left-handedness; (v) inability to undergo any measurements; and (vi) maximum distance amplitude ≥ 30 cm in the bimanual coordination task (20). After applying these exclusion criteria, the remaining 312 participants were included in the analysis (Supplementary Figure 1).

### 2.1 Ethics declarations

This study was conducted in accordance with the Declaration of Helsinki and informed consent was obtained from all participants. This study was approved by the Kyoto Tachibana University Research Ethics Committee (approval number 24-30). This study was registered in the UMIN Clinical Trials Registry (UMIN000056340).

### 2.2 Measures

First, we assessed the frailty of the participants using the Kihon Checklist (KCL). The KCL is a questionnaire developed in Japan to identify older adults who are at high risk of needing care in the near future (21,22). In recent years, the KCL has been used as a tool for frailty assessment (23) and has been recommended as a validated tool in international clinical guidelines for frailty (24,25). The KCL is a self-administered questionnaire with “yes/no” responses that consists of 25 questions covering seven domains: activities of daily living, physical function, nutritional status, oral function, social withdrawal, cognitive function, and depressive mood. In the KCL, higher scores indicate a greater risk of needing care in daily life. In this study, participants with scores of 0–3, 4–7, and ≥8 were categorized into robust, pre-frail, and frail groups, respectively (23).

Next, all the participants performed a bimanual coordination task. Participants sat on chairs with backrests and placed their forearms on a platform. During each task, the forearms were positioned in neutral rotation with the third, fourth, and fifth fingers slightly flexed, and the participants underwent measurements with their eyes closed (Supplementary Figure 2). The bimanual coordination task consisted of two tasks: the in-phase task, wherein tapping movements of the thumb and index finger were performed simultaneously as quickly as possible with both hands; and the anti-phase task, wherein tapping movements alternated between the left and right hands (Supplementary Figure 3) (26,27). The participants performed the 15-s measurement after a 15-s pre-practice in each task. We instructed the participants to “perform as fast as possible and in the same rhythm.”

Finger movements during bimanual coordination tasks were measured using a magnetic sensor finger-tapping device (UB-2, Maxell Ltd. Tokyo, Japan) (26). Magnetic sensors were attached to the participant’s thumb and index finger with a rubber band, and the distance was calculated on the basis of the strength of the magnetic field generated between the two fingers. This device yields highly reproducible and reliable measurements across periods, devices, and measurement examiners (27). During the bimanual coordination task, the participants were instructed to open their fingers to a width of 4 cm to minimize amplitude variations across participants (28,29). The parameters of the bimanual coordination task (distance, tap interval, and phase difference) were obtained from the recorded data (Table 1) (27). Four parameters of "Distance" were used to evaluate the distance and movement amplitude of the thumb and index finger during the task; four parameters of "Tap interval" were used to evaluate the average speed of movement and variability of tapping; and one parameter of "Phase difference" was used to evaluate the timing discrepancy of tapping between the hands.

**Table 1.** Characteristics of the bimanual coordinated task.

|  | Parameter | Description | Assessment |
| --- | --- | --- | --- |
| Distance | Total travel distance (mm) | The sum of the distances moved by the thumb and index finger. The overall amount of movement. | Higher values indicate higher finger dexterity. |
|  | Ave of local max distance (mm) | Average amplitude of the distance waveform. | Values closer to 40 mm indicate higher finger dexterity. |
|  | SD of local max distance (mm) | Variation in the amplitude of the distance waveform. | Lower values indicate higher finger dexterity. |
|  | Slope of approximate line of local max points (mm/s) | The slope is a linear regression of the relationship between the maximum point of each tap and time. As the tap amplitude decreases due to fatigue, the slope increases in the negative direction. When there is no effect of fatigue, the slope is 0. | Lower values indicate higher finger dexterity. |
| Tap interval | Number of taps | Number of taps during the measurement time. | Lower values indicate higher finger dexterity. |
|  | Ave of tap intervals (s) | Average in time difference between two consecutive taps. | Lower values indicate higher finger dexterity. |
|  | Frequency of taps (Hz) | Inverse to the mean of the tap interval. | Higher values indicate higher finger dexterity. |
|  | SD of inter-tap interval (s) | Variations in time difference between two consecutive taps. | Lower values indicate higher finger dexterity. |
| Phase difference | SD of phase difference (degree) | Assuming the interval between one tap is 360°, the time lag between the left and right hands is expressed as an angle. This parameter is the variation of its value. | Lower values indicate higher finger dexterity. |

Ave, average; Max, maximum; SD, standard deviation.

### 2.3 Statistical analysis

Participants were divided into frail, pre-frail, and robust groups based on the KCL results. First, a chi-square test was conducted to compare the male/female ratios among the groups. Participants’ age, height, weight, and MMSE and KCL scores were compared between the groups using one-way analysis of variance (ANOVA). Next, three-way ANOVA with a mixed design was conducted to compare the total travel distance, average of local maximum distance, standard deviation (SD) of local maximum distance, slope of the approximate line of local maximum points, number of taps, average of tap intervals, frequency of taps, and SD of inter-tap interval during the bimanual coordination task, considering hand (left, right), task (in-phase task, anti-phase task), and group (frail, pre-frail, robust) as factors. Additionally, a two-way ANOVA with a mixed design was used to compare the SD of the phase difference between left-and right-hand tapping (SD of phase difference), considering task (in-phase task, anti-phase task) and group (frail, pre-frail, robust) as factors. Bonferroni post-hoc tests were performed for parameters showing significant interactions or main effects in all ANOVAs. Statistical analyses were performed using SPSS version 29.0 (IBM, Armonk, NY, USA), with the significance level set at 5%.

## 3 Results

### 3.1 Characteristics of the participants

Based on the KCL’s assessment of frailty, we divided the participants into the frail (47 participants; 8 males, 39 females), pre-frail (136 participants; 27 males, 109 females), and robust (129 participants; 33 males, 96 females) groups. The results of the chi-square test showed no significant differences among male/female ratios in each group (χ^2^ = 2.10, p = 0.37). One-way ANOVA revealed no significant intergroup differences in age, height, weight, or MMSE score (p > 0.05), but showed significant differences in the KCL score (p < 0.05). Post-hoc tests showed that the KCL scores in the pre-frail and frail groups were significantly higher than that in the robust group, and the score in the frail group was significantly higher than that in the pre-frail group (p < 0.05; Table 2).

**Table 2.**
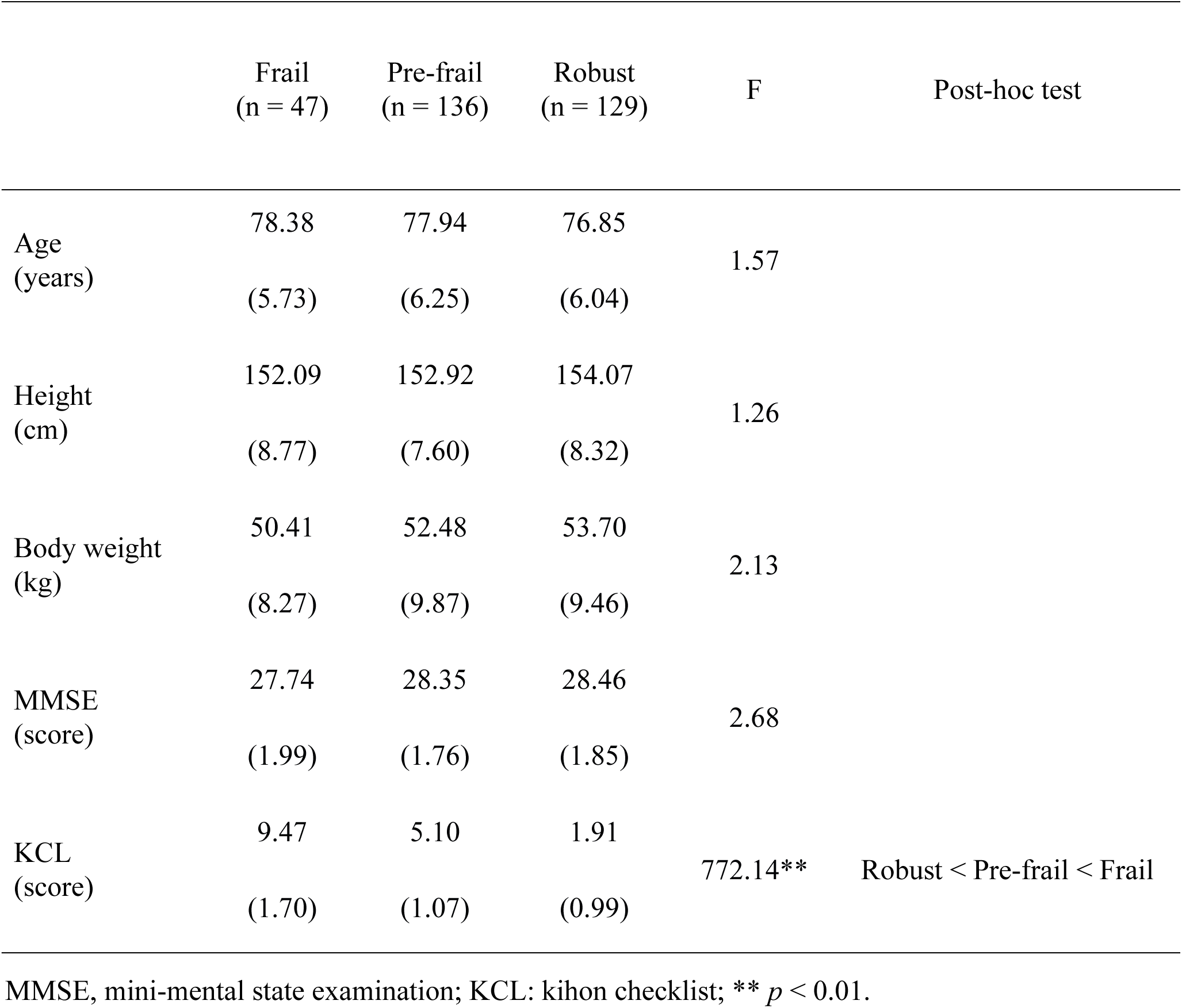
Characteristics of the participants.

|  | Frail<br>(n = 47) | Pre-frail<br>(n = 136) | Robust<br>(n = 129) | F | Post-hoc test |
| --- | --- | --- | --- | --- | --- |
| Age<br>(years) | 78.38<br>(5.73) | 77.94<br>(6.25) | 76.85<br>(6.04) | 1.57 |  |
| Height<br>(cm) | 152.09<br>(8.77) | 152.92<br>(7.60) | 154.07<br>(8.32) | 1.26 |  |
| Body weight<br>(kg) | 50.41<br>(8.27) | 52.48<br>(9.87) | 53.70<br>(9.46) | 2.13 |  |
| MMSE<br>(score) | 27.74<br>(1.99) | 28.35<br>(1.76) | 28.46<br>(1.85) | 2.68 |  |
| KCL<br>(score) | 9.47<br>(1.70) | 5.10<br>(1.07) | 1.91<br>(0.99) | 772.14** | Robust < Pre-frail < Frail |

MMSE, mini-mental state examination; KCL: kihon checklist; \*\* $p < 0.01$ .

### 3.2 Results of three-way ANOVA with hand, task, and group as factors

The three-way ANOVA results showed no significant interactions among the three factors (hand × task × group) for the total travel distance, average of local maximum distance, SD of local maximum distance, slope of the approximate line of local maximum points, number of taps, average of tap intervals, frequency of taps, and SD of the inter-tap interval (p > 0.05). Additionally, no significant hand × group and task × group interactions were observed. Conversely, the slope of the approximate line of local maximum points and the SD of the inter-tap interval showed significant hand × task interactions (p < 0.05). Post-hoc test results indicated that in the in-phase task, the slope of the approximate line of the local maximum points was significantly higher in the right hand than in the left hand (p < 0.05). Additionally, the slope of the approximate line of the local maximum points in the right hand was significantly higher in the in-phase task than in the anti-phase task (p < 0.05). The SD of the inter-tap interval was significantly higher in the left hand than in the right hand in both the in-phase and anti-phase tasks (p < 0.05). In addition, the SD of the inter-tap interval was significantly higher in the anti-phase task than in the in-phase task for both the left and right hands (p < 0.05).

The total travel distance showed a significant main effect of the group factor (p < 0.05). Post-hoc test results indicated that the total travel distance was significantly longer in the robust group than in the frail group (p < 0.05). The total travel distance and average of the local maximum distance, SD of the local maximum distance, slope of the approximate line of local maximum points, number of taps, average of tap intervals, frequency of taps, and SD of the inter-tap interval had significant main effects of the task factor (p < 0.05). The post-hoc test results showed that the total travel distance, number of taps, and frequency of taps showed significant main effects of the task factor (p < 0.05). The frequency of taps was significantly higher in the in-phase task than in the anti-phase task (p < 0.05). The average of the local maximum distance, SD of the local maximum distance, and average of the tap intervals were significantly higher in the anti-phase task than in the in-phase task (p < 0.05). The total travel distance, SD of the local maximum distance, number of taps, average tap interval, frequency of taps, and SD of the inter-tap interval showed significant main effects of the hand factor (p < 0.05). According to the post-hoc tests, the total travel distance, number of taps, and frequency of taps were significantly higher for the right hand than for the left hand (p < 0.05). In contrast, the SD of the local maximum distance and average of tap intervals were significantly higher for the left hand than for the right hand (p < 0.05; Table 3).

**Table 3.**
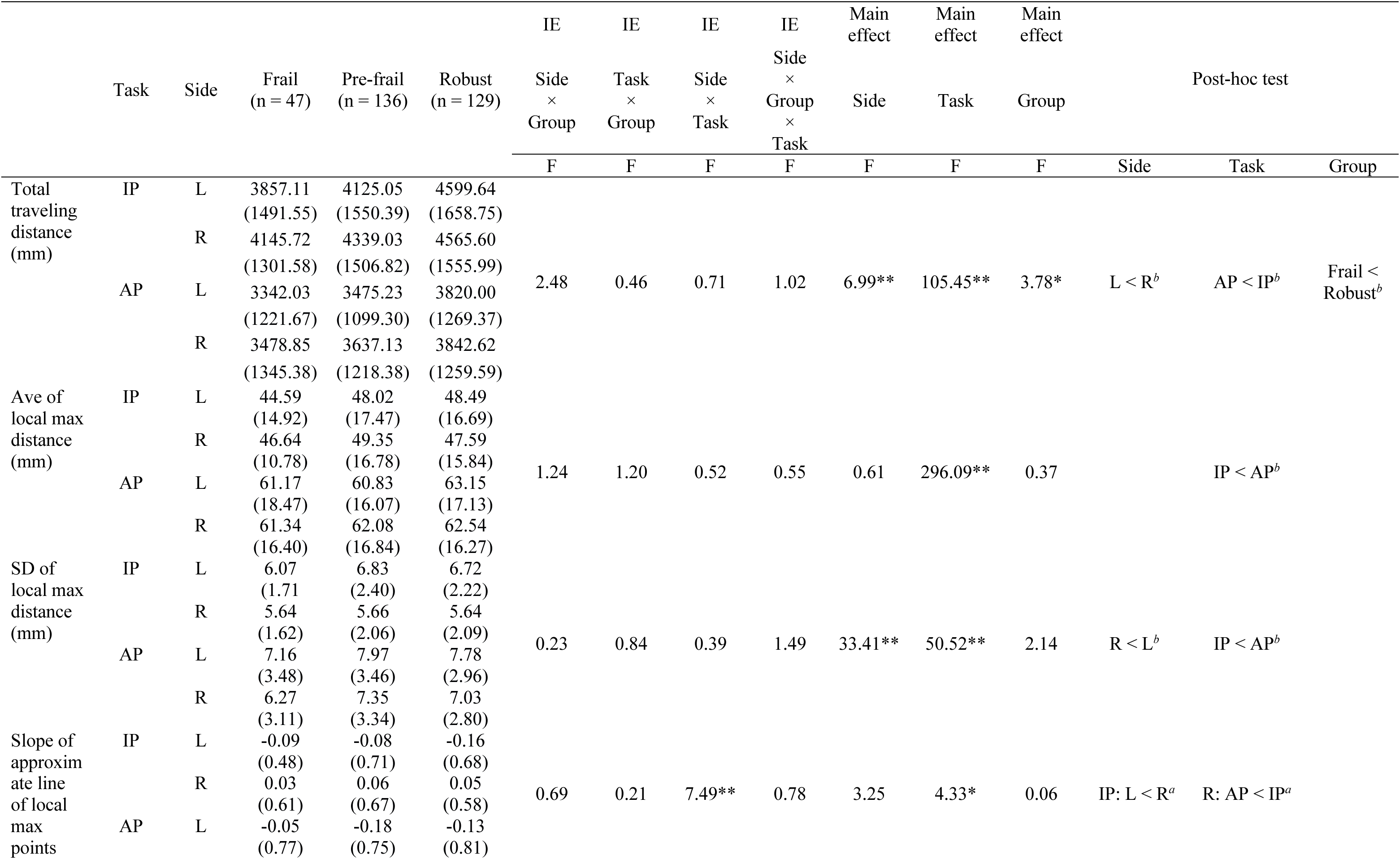
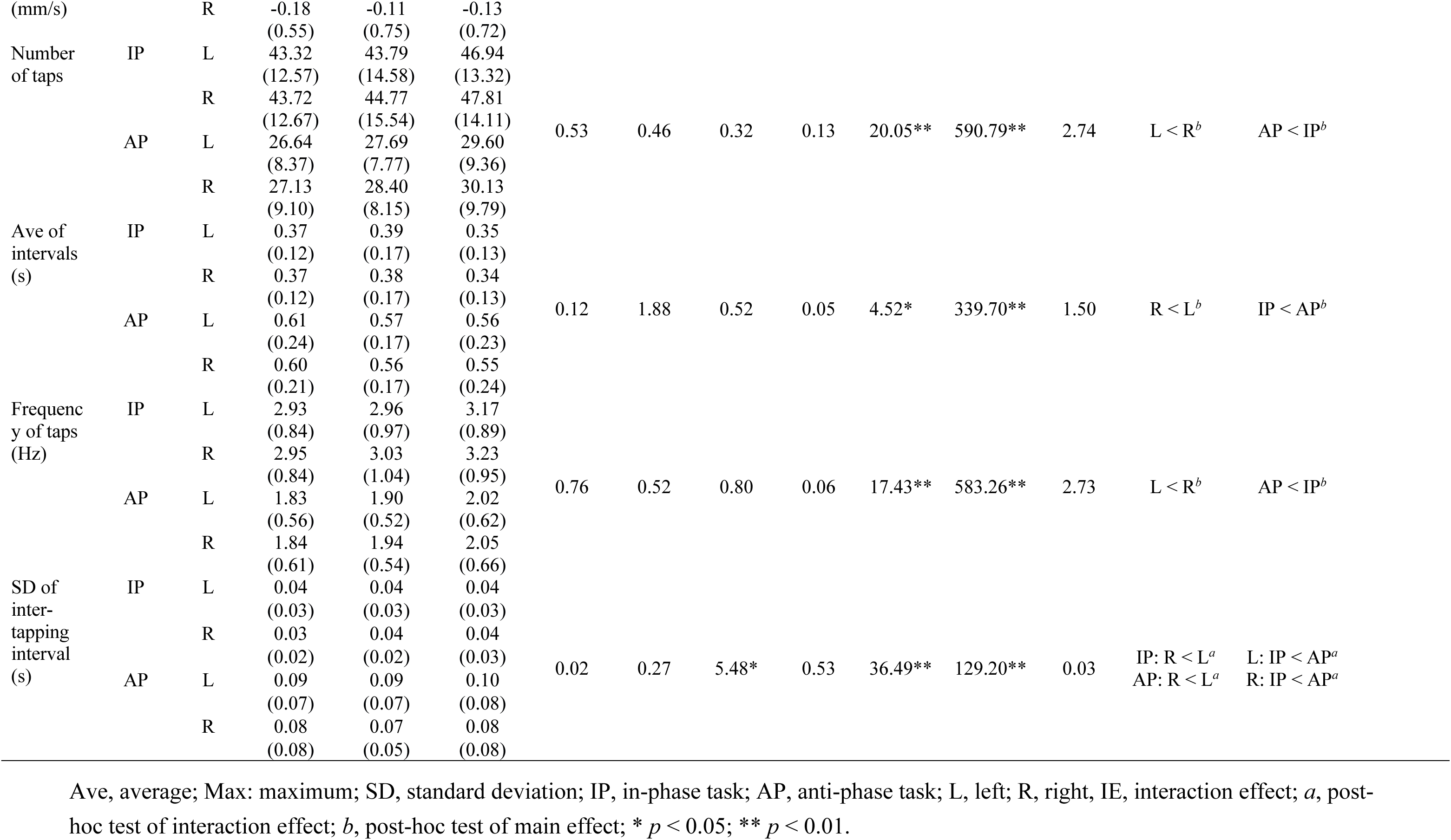
Results of three-way ANOVA with hand, task, and group as factors.

### 3.3 Results of two-way ANOVA with task and group as factors

In the two-way ANOVA, the SD of the phase difference showed no significant interaction between the task and group factors, nor a main effect of the group factors. However, a significant main effect of the task factor was observed (p < 0.05). The post-hoc test results showed that the SD of the phase difference was significantly higher for the anti-phase task than for the in-phase task (p < 0.05; Table 4).

**Table 4.**
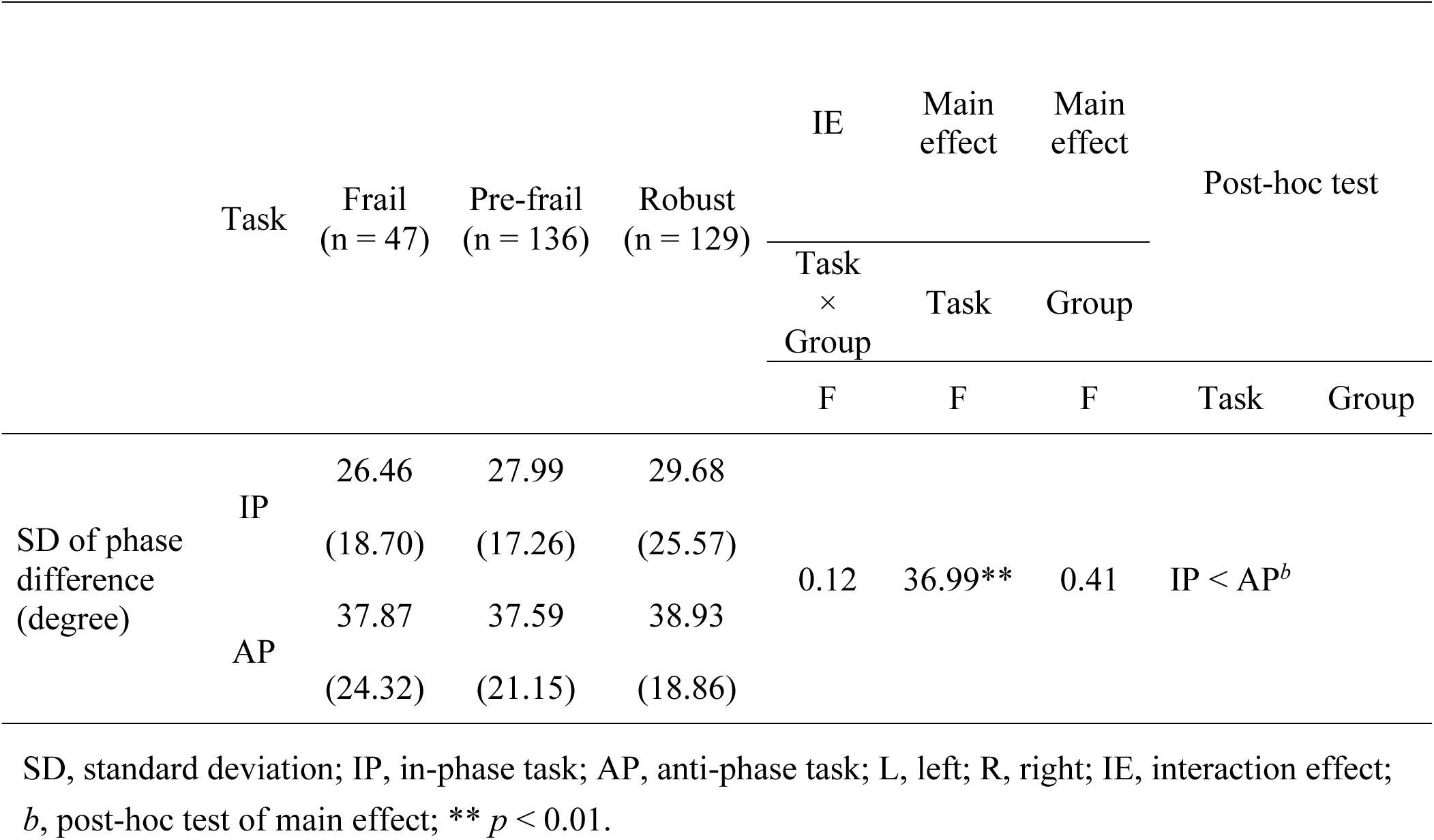
Results of two-way ANOVA with task and group as factors.

## 4 Discussion

This study compared the characteristics of bimanual coordinated movements in community-dwelling older adults with frailty, pre-frailty, and robust health. The results showed that the total distance of the bimanual coordinated movements was shorter in the frail group than in the robust group. Additionally, regardless of the degree of frailty, finger dexterity during the bimanual coordination task was lower in the anti-phase task than in the in-phase task and lower in the left hand than in the right hand. These results suggest that older adults with and without frailty exhibit similar levels of bimanual coordination. However, the amount of movement in bimanual coordination tasks was lower in older adults with frailty than in robust older adults.

### 4.1 Relationship between bimanual coordination and frailty

In this study, older adults were divided into pre-frail, frail, and robust groups and asked to perform a bimanual coordination task consisting of in-phase and anti-phase tasks. The results showed that the total travel distance was shorter in the frail group than in the robust group. The total travel distance is influenced by the velocity and number of movement taps (30), which represents the amount of finger movement. In a study evaluating the relationship between frailty and finger movement control while performing unilateral movements using the dominant hand, agility, smoothness of movement, and strength were reported to be lower in older adults with frailty than in healthy older adults (13). Frailty is defined as “a medical syndrome caused by multiple causes and triggers, characterized by a decline in muscle strength and endurance and a decrease in physiological function, with an increased vulnerability to requiring care or death” (5). Since a decrease in muscle strength leads to a decline in movement velocity (31), the frail group in this study may have had slower bimanual coordinated movements than the robust group, resulting in the shorter total travel distance in the frail group than in the robust group. Although the difference in the amount of movement between the frail and robust groups could also be attributed to reduced endurance in the frail group (5,32), the slope of the approximate line of the local maximum points, which reflects the effect of fatigue based on the relationship between the maximum distance between two fingers per tap and time, showed no significant difference between the frail and robust groups in this study. Therefore, we inferred that a decline in endurance was unlikely to affect the amount of movement in the bimanual coordinated movement task. Interhemispheric interactions via the corpus callosum have also been reported to play an important role in bimanual coordinated movements (33). The structural and functional connectivity of the corpus callosum has been shown to be altered with age, leading to reduced performance in bimanual coordinated movements (34,35). Furthermore, structural alterations in the cerebral corpus callosum have been reported to be more pronounced in older adults with frailty than in healthy older adults (36). Therefore, the frail group may have had reduced finger dexterity due to altered structural and functional connectivity of the cerebral corpus callosum, which may have caused the differences in performance of the bimanual coordination task between the robust and frail groups.

### 4.2 Comparison of in-phase and anti-phase tasks

Our findings also showed that the total travel distance, number of taps, and frequency of taps were higher in the in-phase task than in the anti-phase task. The average of the local maximum distance, SD of the local maximum distance, average of the tap intervals, and SD of the phase difference were higher in the anti-phase task than in the in-phase task. The slope of the approximate line of the local maximum points is suspected to be influenced by fatigue because the distance between the two fingers becomes narrower over time if the slope has a negative value. In the present study, the slope of the approximate line of the local maximum points was negative for the anti-phase task and positive for the in-phase task for the right hand. Therefore, the anti-phase task may have been affected by fatigue. In addition, the SD of the inter-tap interval showed that the rhythm of movement was more variable in the anti-phase task. The number of taps, average tap interval, and frequency of taps indicated that the anti-phase task involved fewer taps, a lower frequency, and longer periods than the in-phase task. The total travel distance and the average and SD of the local maximum distance revealed that the amount of movement was smaller in the anti-phase task than in the in-phase task and that the distance between two fingers per tap and its variation were larger in the anti-phase task than in the in-phase task. The SD of the phase difference showed that the anti-phase task had more timing deviations than the in-phase task for both tasks. Thus, in this study, the anti-phase task was inferior to the in-phase task for all parameters of finger dexterity. The anti-phase task requires specific muscle activity with continuous timing to maintain alternating bimanual movements, and this timing is asymmetric between the left and right hands (37). In addition, maintaining attention is necessary to preserve the phase relationship between hands. For anti-phase tasks and cognitive function, research involving community-dwelling older adults with declining cognitive function has shown a correlation between tapping velocity in the anti-phase task and decline in working memory and attention (29). Therefore, the anti-phase task, which requires independent alternating movements of both hands, is suggested to be more challenging than the in-phase task or unilateral motor tasks and is prone to differences in finger function (28). Thus, similar to robust older adults, older adults with frailty in this study may have experienced higher difficulty in the anti-phase task than in the in-phase task and showed characteristics of reduced finger dexterity for each parameter.

### 4.3 Comparison of left and right hand in bimanual coordinated movement

Since the participants performed the same finger-tapping task with their left and right hands, we expected no significant differences between the parameters for each hand. However, the total travel distance was significantly longer with the right hand than with the left. In addition, the SD of the local maximum distance was lower for the right hand than for the left hand. The right hand showed a higher number, and frequency, of taps as well as longer intervals than the left hand. Furthermore, the SD of the inter-tap interval was smaller for the right hand than for the left hand. If the thumb is repeatedly moved in a specific direction, the trained movement increases cortical excitability (38). Thus, repetitive movements induces plastic reorganization in the primary motor cortex, and this phenomenon is called use-dependent plasticity (39,40). This use-dependent plasticity has been found to inhibit motor errors and reduce motor planning time, even in complex daily activities (41). The dominant hand is used more frequently than the non-dominant hand in daily life, and older adults are trained to use the dominant hand in their daily activities themselves (34). These findings suggest that the primary motor cortex innervating the dominant hand enables spatially and temporally efficient movements through use-dependent plasticity (42). In the present study, the right hand may have had higher finger dexterity than the left hand for all parameters, regardless of other factors. Therefore, older adults with frailty, like robust older adults, have higher finger dexterity during bimanual coordination tasks with their right hand than with their left hand.

### 4.4 Limitations

The limitations of this study include the fact that it examined the characteristics of bimanual coordination only at the behavioral level and did not examine the neural mechanisms underlying bimanual coordination. Previous studies comparing unilateral movements in participants of a wide range of ages, from children to healthy older adults, have shown that immature or degenerated motor systems may maintain or improve performance by bilateral mobilization of brain regions in comparison with normal motor systems (35,43). Since structural changes in the corpus callosum have been shown to occur in older adults with frailty (28), older adults with and without frailty may have shown differences in the neural mechanisms in the brain during the bimanual coordination task in this study. Future studies should examine interhemispheric inhibition and facilitation functions during bimanual coordination tasks using techniques such as transcranial magnetic stimulation to elucidate the neural mechanisms underlying bimanual coordination in older adults with frailty.

## 5 Conclusions

This study characterized bimanual coordinated movements in older adults with frailty, pre-frailty, and robust health. Based on the bimanual coordination task, the total traveling distance was shorter in the frail group than in the robust group. Regardless of the severity of frailty, participants showed lower bimanual coordination in the anti-phase task than in the in-phase task, and finger dexterity during the bimanual coordination tasks was lower in the left hand than in the right hand. Thus, while older adults with frailty exhibit bimanual coordination similar to that of robust older adults, the amount of movement in the bimanual coordination task by those with frailty is lower than that by robust older adults. The results of this study suggest that bimanual coordination tasks may be applicable as an assessment tool for frailty.

## 7 Author Contributions

Author contributions included conceptualization (SM, and HN), data curation (SF), formal analysis (SF), funding acquisition (SM, TM, KN, and HN), investigation (SF, SM, AG, SS, YS, and HN), methodology (SM, and HN), project administration (SM, and HN), resources (SM, and HN), supervision (SM, and HN), visualization (SF), writing – original draft (SF), writing – review & editing (SF, SM, AG, SS, YS, TM, KN and HN), and approval of final version to be published and agreement to be accountable for the integrity and accuracy of all aspects of the work (all authors).

## Acknowledgments

We would like to express our sincere gratitude to all participants for their willingness to participate in this study. We gratefully acknowledge Mr. Tomohiko Mizuguchi at Maxell, Ltd. for advice and suggestions of analysis.

## 11 Data Availability Statement

The data that support the findings of this study are available on request from the corresponding author, HN, upon reasonable request. The data are not publicly available due to their containing information that could compromise the privacy of research participants.

