## Supplementary Figure for "Comparison of characteristics of bimanual coordinated movements in older adults with frailty, pre-frailty, and robust health"

### *Supplementary Material*

#### 1 Supplementary Figures

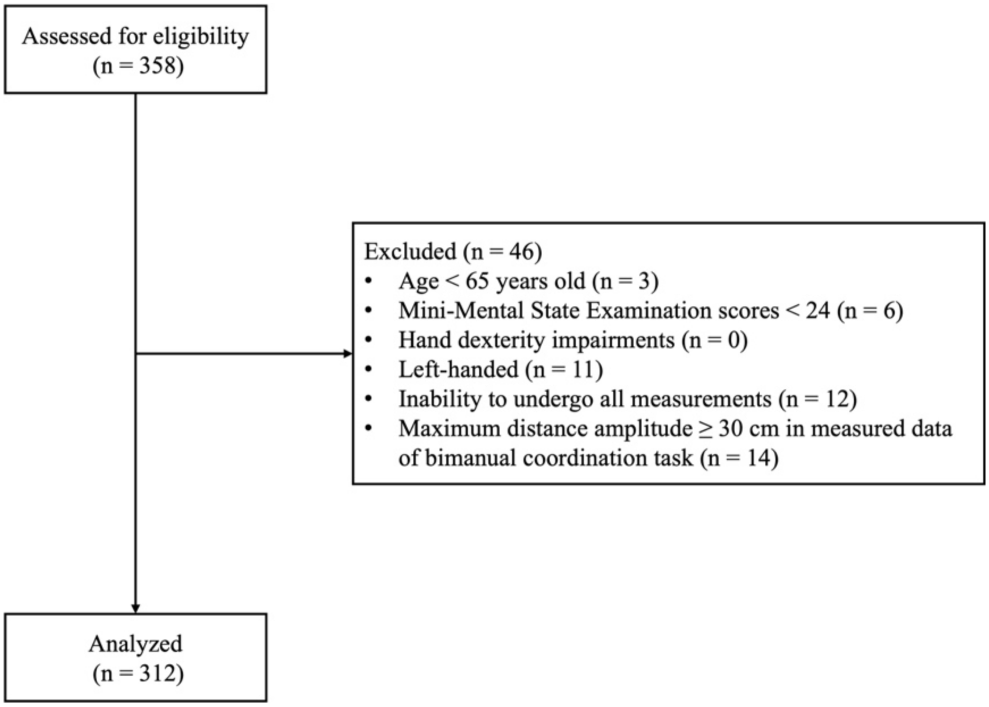


**Supplementary Figure 1. Flowchart of participant selection.** Participants with age < 65 years, Mini-Mental State Examination scores < 24, hand dexterity impairments due to musculoskeletal or central nervous disease, left-handedness, inability to undergo all measurements, and maximum distance amplitude ≥ 30 cm in the measured data of the bimanual coordination task were excluded. Finally, this study included 312 participants.

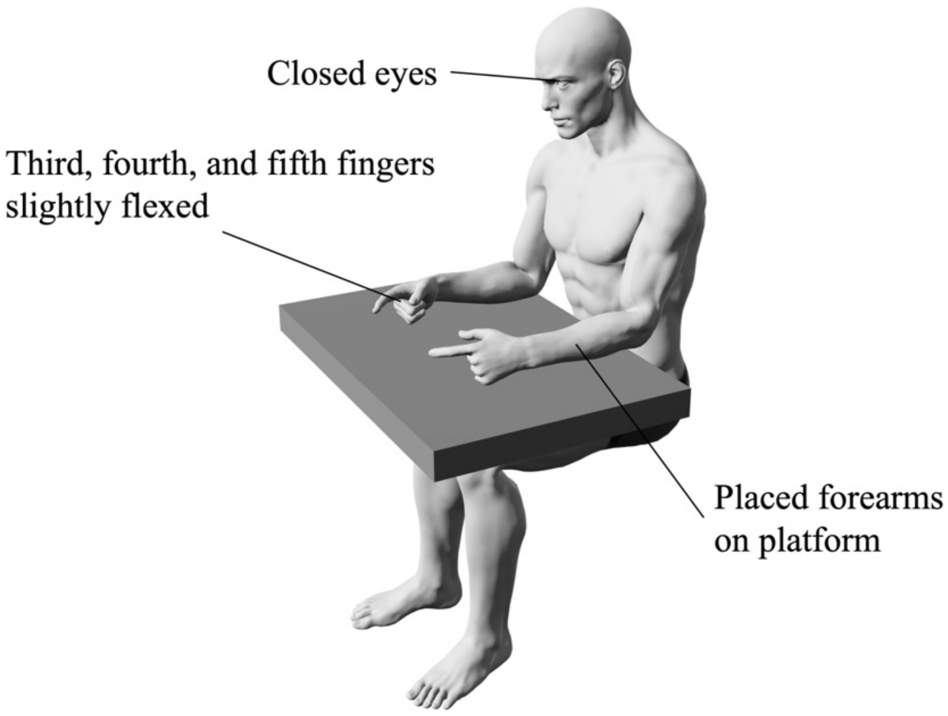


**Supplementary Figure 2. Measurement position in the bimanual coordination task.** Participants sat on chairs with backrests and placed their forearms on the platform. During each task, the forearms were positioned in neutral rotation, with the third, fourth, and fifth fingers slightly flexed, and participants underwent measurements with closed eyes.

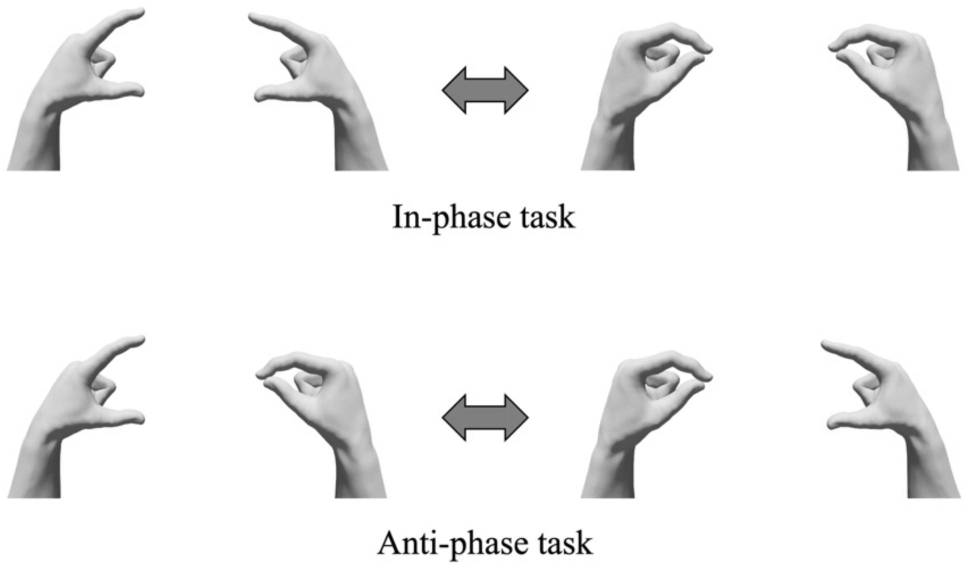


**Supplementary Figure 3. Bimanual coordination task.** (A) In-phase task: Participants performed tapping movements of the thumb and index finger simultaneously on both sides. (B) Anti-phase task: Participants performed tapping movements of the thumb and index finger alternately on both sides.
